# Racial and ethnic inequalities in COVID-19 mortality within Texas carceral settings

**DOI:** 10.1101/2021.09.26.21264145

**Authors:** Neal Marquez, Destiny Moreno, Amanda Klonsky, Sharon Dolovich

## Abstract

Several analyses have highlighted racial and ethnic disparities related to COVID-19 health outcomes across the United States. Less focus has been placed on more localized contexts, such as carceral settings, where racial and ethnic inequities in COVID-19 health outcomes also exist, but the proximal drivers of inequality are different. In this study, we analyzed mortality rates among incarcerated people in the Texas Department of Criminal Justice (TDCJ) to assess racial and ethnic differences in COVID-19 mortality. We obtained monthly demographic and mortality information of the TDCJ population from April 1, 2019 to March 31, 2021 from TDCJ monthly reports and open record requests filed by the Texas Justice Initiative. We estimated the risk of COVID-19 mortality for the Hispanic and Black population relative to the White population using a Bayesian regression framework, adjusting for sex and age. In the first 12 months of the pandemic, Hispanic and Black all-cause mortality rates were higher than that of the White population, reversing the pattern observed the 12 months prior. Adjusted risk of COVID-19 mortality relative to the White population was 1.96 (CI 1.32–2.93) for the Hispanic population and 1.66 (CI 1.10–2.52) for the Black population. We find that COVID-19 mortality has disproportionately impacted Hispanic and Black individuals within the TDCJ population. As the proximal mechanisms which drive these inequalities are likely different than those which lead to racial inequalities in the non-incarcerated populations, future studies should look to assess and address the specific drivers of COVID-19 related disparities in carceral settings.

## BACKGROUND

Racial and ethnic disparities related to COVID-19 morbidity and mortality in the United States have been well known since early in the pandemic (Mackey et al., 2021). It is widely agreed that socio-economic factors — which some refer to as the social determinants of health — drive these disparities, as opposed to underlying biological differences (Mackey et al., 2021; Yehia et al., 2020). Although several studies have highlighted major social factors driving national-level inequalities in COVID-19 related health outcomes, social determinants of health impact communities at multiple levels (Mackey et al., 2021; Selden and Berdahl, 2020; Millett et al., 2020) and manifestations of systemic racism have been found to generate pervasive health inequalities (Barten et al., 2007; Boyd et al., 2020). Understanding how social, economic, and political inequalities operate at local levels and in specific communities are essential for creating more equitable health outcomes (Barten et al., 2007).

In the context of COVID-19, carceral systems have appeared to play a critical role in exacerbating health inequalities. Incarcerated people have both a COVID-19 incidence and a mortality rate several times greater than the U.S.population as a whole (Saloner et al., 2020). As is well known, the incarcerated population in the U.S. is disproportionately Black and Hispanic (Western and Pettit, 2010). Consequently, these individuals have been assumed to be disproportionately impacted by the high risk of COVID-19 within carceral settings (Nowotny et al., 2021). Furthermore, communities with high rates of incarceration have been shown to suffer adverse health effects from high concentrations of COVID in prisons and jails, as people exposed to COVID-19 inside return to their communities once released (Reinhart and Chen, 2021).

While differential rates of incarceration among racial and ethnic groups have been shown to exacerbate racial and ethnic inequalities in COVID-19 burden in the United States as a whole, inequalities are likely to be present within carceral institutions as well. Racial disparities have been documented throughout the criminal legal system, contributing to racial and ethnic differences at multiple points in the process, including police interaction, bail decisions, sentencing, parole, and the application of the “collateral consequences” of a criminal conviction (The Sentencing Project, 2018). While some studies have found that racial and ethnic health inequalities are somewhat lessened in carceral contexts (Nowotny et al., 2017), it is well established that the COVID-19 pandemic exacerbated racial and ethnic health inequalities in the U.S. population as a whole (Andrasfay and Goldman, 2021). Several factors at work in the prison context, including health provider barriers, financial concerns, a lack of institutional trust, crowded living arrangements, and a high prevalence of pre-existing conditions, make it likely that the same is true in carceral settings.

To assess the degree to which incarceration contributes to racial and ethnic inequalities in COVID-19 related-health outcomes, we examine COVID-19 mortality outcomes within the Texas Department of Criminal Justice (TDCJ). In addition to managing several non-prison facilties, TDCJ holds more people in prisons than any other state Department of Corrections in the U.S. Furthermore, TDCJ holds the largest number of Black and Hispanic individuals in prison settings. Black and Hispanic people comprise 66.3% of the TDCJ prison population, despite only representing 50.1% of the state’s total population (authors calculations). Using roster data for the TDCJ population and mortality data obtained through monthly open records requests filed by the Texas Justice Initiative, we investigate whether COVID-19 altered patterns of mortality for Black, Hispanic, and White individuals within the TDCJ population, and if so, which groups were most heavily impacted.

## RESULTS

During the study period, April 1, 2019 to March 31, 2021, the median ages within the Black, Hispanic, and White populations were 37, 36, and 39 respectively. The shares of the population over the age of 65, the age group found to be most vulnerable to COVID-19 related mortality, were 2.49% for the Black population, 2.34% for the Hispanic population, and 4.62% for the White population. During the first 12 months of the pandemic (pandemic period, April 1, 2020 to March 31, 2021), the White population had the highest number of total deaths (283) while the Hispanic population had the largest number of COVID-19 related deaths (104) (Table 1).

**Table 1.** Demographic and mortality statistics of TDCJ population. Population statistics computed over the 24 month period of the study. Mortality rates are sex and age standardized per 10,000 person-years.

|  | Black | Hispanic | White | Total |
| --- | --- | --- | --- | --- |
| **Demographics** |  |  |  |  |
| Average Monthly Population | 43,602 | 43,868 | 44,403 | 131,873 |
| Percent 65+ | 2.49% | 2.34% | 4.62% | 3.16% |
| Percent Male | 94.37% | 94.56% | 87.58% | 92.15% |
| **Standardized Mortality Rate (Deaths)** |  |  |  |  |
| All-Cause Mortality<br>*Pre-Pandemic Period* | 28.77<br>(122) | 31.24<br>(122) | 35.77<br>(210) | 32.49<br>(454) |
| All-Cause Mortality<br>*Pandemic Period* | 64.89<br>(237) | 64.75<br>(213) | 54.31<br>(283) | 60.06<br>(733) |
| COVID-19 Mortality<br>*Pandemic Period* | 25.41<br>(93) | 33.06<br>(104) | 16.35<br>(91) | 23.61<br>(288) |

During the pandemic period, the standardized all-cause mortality rate across all three groups had increased by 85% over the previous 12 months (pre-pandemic period). Table 1 shows the standardized all-cause mortality rates of Black, Hispanic, and White populations both for the pre-pandemic as well as the pandemic period. The increase was greatest for the Black population, which saw a 126% increase, followed by the Hispanic population (107% increase), and was lowest for the White population (52% increase). In the pre-period, the White population had the highest standardized all-cause mortality rate. For the pandemic period, these patterns reversed, with the White population having a lower standardized all-cause mortality rate than either the Black or Hispanic populations (Table 1).

Standardized COVID-19 mortality rates were 25.41 for the Black population, 33.06 for the Hispanic population, and 16.35 for the White population (Table 1). The Black population had a standardized COVID-19 mortality rate which was 1.6 times greater than the White population, while the Hispanic population standardized COVID-19 mortality rate was 2.0 times greater than the White population.

Adjusting for age and sex in a Bayesian regression framework, our analysis finds that, relative to the White population, risk of COVID-19 related mortality during the pandemic period for Blacks was 1.66 (CI 1.10-2.52) while for Hispanics it was 1.96 (CI 1.32–2.93). During the same time period, relative to the White population, risk of all-cause mortality was 1.19 (CI 0.97-1.44) for the Black population and 1.17 (CI 0.95-1.44) for the Hispanic population (Table 2).

**Table 2.** Bayesian Model Incidence Rate Ratios. Model results for Bayesian negative binomial regression. 95% credible intervals of relative risk estimates shown in parenthesis. Individuals aged 20-34, who were White, and female were used as the baseline category. Full model specifications can be found in the SI appendix.

| Predictors | COVID-19 Mortality<br>*Relative Risk* | All-Cause Mortality<br>*Relative Risk* |
| --- | --- | --- |
| Age: 35-44 | 5.69<br>(1.89 – 21.88) | 1.97<br>(1.32 – 2.91) |
| Age: 45-54 | 29.73<br>(10.84 – 107.50) | 5.84<br>(4.12 – 8.34) |
| Age: 55-64 | 98.80<br>(37.38 – 352.19) | 17.11<br>(12.29 – 24.14) |
| Age: 65-74 | 388.45<br>(143.28 – 1394.84) | 52.80<br>(37.72 – 75.67) |
| Age: 75+ | 784.86<br>(269.64 – 2973.62) | 124.23<br>(83.91 – 186.13) |
| Race: Black | 1.66<br>(1.10 – 2.52) | 1.19<br>(0.97 – 1.47) |
| Race: Hispanic | 1.96<br>(1.32 – 2.93) | 1.17<br>(0.95 – 1.44) |
| Sex: Male | 1.10<br>(0.59–2.19) | 1.76<br>(1.17 – 2.81) |
| Observations | 429 | 429 |

## DISCUSSION

Although all individuals in TDCJ carceral settings are supposed to have similar housing and access to constitutionally mandated health care, our analysis finds that, during the first twelve months of the pandemic, Black and Hispanic populations experienced higher rates and risk of standardized COVID-19 mortality when compared to their White counterparts. Over this time period, the risk of COVID-19 mortality was 1.96 times higher for the Hispanic population than for the White population, and 1.66 times as high for Black population as for the White population in TDCJ facilities. Moreover, the standardized COVID-19 mortality rate among the Hispanic population during the pandemic period was higher than the rate of standardized all-cause mortality in the pre-pandemic period. In the pandemic period, the Hispanic population in TDCJ custody had a higher standardized COVID-19 mortality rate than standardized non-COVID-19 mortality rate.

These disparate outcomes suggest that individuals in TDCJ custody are subject to different experiences during incarceration, stratified by race and ethnicity, with inequitable levels of either COVID-19 risk, COVID-19 exposure, or barriers to health care utilization to prevent COVID-19 related mortality. Given the continued elevation of COVID-19 infection rates due to Delta and other variants, the likelihood of a future pandemic, and the high risk of infectious disease spread in carceral settings (National Academies of Sciences, 2020), it is essential both to protect the health of incarcerated populations more generally and to address the systemic factors that lead to racial and ethnic inequalities of health outcomes in custody.

To be sure, COVID-19 had a substantial impact on patterns of mortality for all three ethnic and racial groups analyzed in this study. The standardized mortality rate across the whole of the TDCJ population increased by 85% from the pre-pandemic to the pandemic period, with COVID-19 related deaths accounting for more than 39% of all deaths.

However, our findings show that the impact COVID-19 had on mortality in TDCJ was meaningfully greater for the Black and Hispanic populations. In addition to COVID-19 mortality being higher, all-cause mortality rates were also higher during the pandemic period for Black and Hispanic populations when compared to the White population, reversing the pattern observed in the 12 months prior to the start of the pandemic. Furthermore, these patterns were not isolated to a particular age group. Every age group in this study had a higher age-specific sex-standardized Hispanic COVID-19 mortality rate than White population, and all but one age group, 65-74, had a higher age-specific sex-standardized Black COVID-19 mortality rate when compared with the COVID-19 mortality rate in the White population.

Examining the causal pathways that led to larger rates of COVID-19 mortality among the Hispanic and Black population is beyond the scope of this study. Still, this and previous analyses provide evidence pointing away from racial disparity in rates of pre-existing conditions as the sole explanatory factor. In a previous study of the TDCJ population, Hispanic individuals were generally found to have a lower age-standardized prevalence of pre-existing conditions compared to the White population, while the Black population had a higher prevalence for some conditions but lower prevalence for others (Harzke et al., 2010). For pre-existing conditions alone to explain the observed differences of this study, a large shift in disease prevalence among the TDCJ population would have had to occurred.

### Implications

If policymakers are to address racial and ethnic inequalities in COVID-19 related health outcomes, understanding which structural factors impact or create these inequalities is essential. The relationship between exposure to carceral settings and increased risk of COVID-19 infection and mortality is well known (Saloner et al., 2020), and greater representation of the Black and Hispanic communities in jails and prisons implies that they are more likely to experience adverse health outcomes related to COVID-19 (Nowotny et al., 2021; Reinhart and Chen, 2021). This study adds to previous research by demonstrating racially inequitable COVID-19 health outcomes within prison settings, a finding suggesting that the structures of systemic racism that lead to racial and ethnic health disparities of COVID-19 in society as a whole, also penetrate the boundaries of carceral systems, as suggested by other scholars (Nowotny et al., 2021). Given that COVID-19 continues to disproportionately impact people in carceral settings, this possibility calls out for further investigation. Yet many state Departments of Corrections have not released information regarding COVID-19 cases and mortality by race or ethnicity (Nowotny et al., 2021).

It is unlikely that the results found here are unique to TDCJ. It is imperative to determine the extent to which the racial inequities related to COVID-19 health outcomes, such as this analysis reveals in the Texas state carceral system, is indeed replicated in other jurisdictions—and steps be taken to identify and address the root causes of existing inequities (National Academies of Sciences, 2020; Nowotny et al., 2021).

Other research has found that COVID-19 exposure and health care access are the most consistent factors explaining racial and ethnic disparities in COVID-19 health outcomes in the United States as a whole (Mackey et al., 2021). Future studies should examine how racial and ethnic groups within carceral settings may be subject to differential COVID-19 exposure risk and the extent to which, due to provider- or system-level barriers to access, racial and ethnic groups may have had differential access to health care within prisons during the pandemic period.

## METHODS

### Data

Monthly population data by age, sex, race, and ethnicity was ascertained from Texas Department of Criminal Justice (TDCJ) high-value data set monthly reports from April 1st, 2019 to March 31st, 2021 (Texas Department of Criminal Justice, 2021). In compliance with Texas Senate Bill 701, TDCJ produces a monthly roster of all individuals in TDCJ custody, along with information about age, sex, and race/ethnicity for all individuals. We aggregated this data to get a count of all individuals by sex, age group, and race/ethnicity for every month of the analysis for the TDCJ custodial population. We were unable to obtain all of the monthly reports TDCJ released during 2019. Where these data were missing, we used linear interpolation to calculate values for a particular population group. As the population of individuals in TDCJ custody who are not Black, Hispanic, or White is less than 0.5% of the total population across the study period, we did not include members of those groups in this analysis.

Mortality data about individuals in TDCJ custody was taken from two data sources constructed by the Texas Justice Initiative (Texas Justice Initiative, 2021). The first source, *Texas Deaths in Custody* is a record of all deaths of individuals in TDCJ custody since 2015. This data set includes date of death, age, sex, and race/ethnicity for each deceased individual listed. The second source, *COVID-19 Deaths in Texas Incarceration Facilities*, is a curated data set of individuals with COVID-19 listed as one of their causes of death. All individuals who appear in the COVID-19 specific data set also appear in the all-cause mortality data set. The Texas Justice Initiative created these data sets by filing open record requests with the Texas Office of the Attorney General for all deaths which occurred in a carceral setting from 2015 onward. As mandated by the Texas Criminal Procedure Code, this information is a matter of public record. We limited our study to include only individuals who died in TDCJ custody and for which full demographic data were available. For four coroner records, one in 2019 and three in 2020, the age of individuals was not provided so these records were removed from the analysis. Because individuals younger than age 20 represent only 1% of the population and because only a single death occurred among this age group during the two years of the study period, we did not include this age group in our analysis.

For each month in the analysis, we used monthly population reports to construct person-months (persons in TDCJ custody during a given month) of population groups in TDCJ by sex (Male, Female), race, ethnicity (Black, Hispanic, White), and age (20-24, 25-34, 35-44, 45-54, 55-64, 65-74, 75+). We divided our analysis into two 12-month periods, April 1, 2019 to March 31, 2020 (pre-pandemic period) and April 1, 2020 to March 31, 2021 (pandemic period). We chose April 2020 as the start of the pandemic period because it was the first month in which a COVID-19 death was reported in TDCJ custody.

### Standardized Mortality Rates

We calculated standardized all-cause and COVID-19 mortality rates by adjusting mortality rates by age and sex to match the population distribution of the total TDCJ population during the pre-pandemic period. Standardized mortality rates were calculated for both the pre-pandemic and pandemic period for both all-cause and COVID-19 mortality and for each racial and ethnic group. Rates are reported per 10,000 person-years.

### Bayesian Regression Models

To model the risk of all-cause and COVID-19 mortality by race/ethnicity we used a Bayesian model framework. Specifically, we modelled age(*a*), race/ethnicity(*r*), sex(*s*) specific monthly(*m*) mortality counts (D) during the pandemic period using a negative binomial distribution where the monthly population (P) for a group is used as an offset. Demographic variables were included as explanatory variables. Because no COVID deaths occurred among individuals younger than 25, for purposes of this analysis we combined age groups 20-24 and 25-34. Individuals aged 20-34, who were White, and female were used as the baseline category (intercept) in these analyses. Model prior and hyper-prior specifications are shown in Equation 1. We modeled all-cause and COVID-19 related mortality separately as outcomes. We have a particular interest in the exponential of the *β* coefficients related to race/ethnicity which denote the relative risk of either COVID-19 or all-cause mortality for either the Black or Hispanic population relative to the White population as shown in Table 2 of the main text.

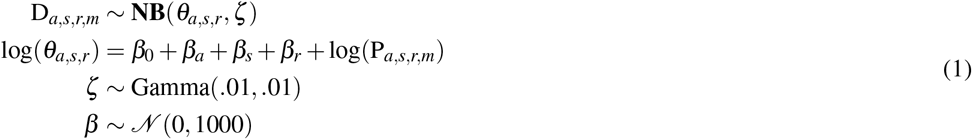

Each model, COVID-19 and all-cause, were fit with a No-U-Turn sampler as implemented in the Stan language using the brms R package as an interface (Bürkner, 2017). Models were fitted with 3 chains and 10,000 iterations, the first 5,000 of which were discarded as burn-in. Model parameter convergence was assessed using the Gelman-Rubin diagnostic with an upper CI limit of 1.1 (Gelman and Rubin, 1992). All parameters were found to converge for both models. All analyses were run in the R programming language version 4.1.1 (R Core Team, 2021). UCLA IRB deemed this study to be exempt.

### Data Availability

Mortality data is made publicly available by the Texas Justice Initiative. Population and demographic data is publicly available by request to TDCJ.

## Data Availability

https://texasjusticeinitiative.org/data

https://www.tdcj.texas.gov/kss_inside.html

## Acknowledgements

The authors would like to thank Michele Deitch and Alycia Welch of the COVID, Corrections, and Oversight Project at the Lyndon B. Johnson School of Public Affairs, University of Texas at Austin, Eva Ruth Moravec from the Texas Justice Initiative, and Amy Harzke from the University of Texas Medical Branch - Galveston for their insight into this analysis. We would also like thank the Texas Justice Initiative as an institution for their efforts to make more accessible information related to health outcomes for those who are justice involved.

## Notes

### Competing Interest Statement

The authors have declared no competing interest.

### Funding Statement

NM, DM, AK, and SD were funded by Arnold Ventures. NM, AK, and SD were funded by Vital Projects Fund. NM was funded by a National Institute of Child Health and Human Development (NICHD) P2C grant (P2C HD042828). SD and AK were funded by a US Centers for Disease Control and Prevention contract.

### Author Declarations

UCLA IRB deemed this study to be exempt.

### Summary of Updates

Table and language fix.

